# Association between Serum Biomarker Profile and Real-World Evidence of Disability in Multiple Sclerosis

**DOI:** 10.1101/2022.10.21.22281364

**Authors:** Wen Zhu, Chenyi Chen, Lili Zhang, Tammy Hoyt, Elizabeth Walker, Shruthi Venkatesh, Fujun Zhang, Ferhan Qureshi, John F. Foley, Zongqi Xia

**Affiliations:** Department of Neurology, University of Pittsburgh, Pittsburgh, PA; Rocky Mountain Multiple Sclerosis Clinic, Salt Lake City, UT; Octave Bioscience, Inc., Menlo Park, CA

**Keywords:** multiple sclerosis, serum, biomarkers, patient-reported outcome, disability, machine learning

## Abstract

**Objective:** Biomarkers could inform disease worsening and severity in people with MS (pwMS). Few studies have examined blood biomarkers informative of patient-reported outcome (PRO) of disability in pwMS. In this study we examine the associations between serum protein biomarker profiles and patient-reported disability in pwMS.

**Methods:** This cross-sectional study included adults with a neurologist-confirmed diagnosis of MS from the University of Pittsburgh Medical Center (Pittsburgh, PA) and the Rocky Mountain MS Clinic (Salt Lake City, Utah) between 2017 and 2020. For exposure, we included 19 serum protein biomarkers potentially associated with MS inflammatory disease activity and 7 key clinical factors (age at sample collection, sex, race/ethnicity, disease subtype, disease duration, disease-modifying treatment, and time interval between sample collection and closest PRO assessment). Using 6 machine learning approaches (Least Absolute Shrinkage and Selection Operator [LASSO] regression, Random Forest [RF], XGBoost, Support-Vector Machines [SVM], stacking ensemble learning, and stacking classification algorithm), we examined model performance in predicting Patient Determined Disease Steps (PDDS) as the primary outcome. We assessed model prediction of Patient-Reported Outcomes Measurement Information System (PROMIS) physical function in a subgroup. We reported model performance using the held-out testing set.

**Results:** We included 431 unique participants (mean age 49 years, 81% women, 94% non-Hispanic White). Using binary outcomes, models comprising both routine clinical factors and the 19 proteins as features consistently outperformed base models (containing clinical features alone or clinical features plus single protein) in predicting severe (PDDS≥4, PROMIS<35) versus mild/moderate (PDDS<4, PROMIS≥35) disability for all machine learning approaches, with LASSO achieving the best area under the curve (AUC_PDDS_=0.91, AUC_PROMIS_=0.90). Using ordinal/continuous outcomes, LASSO models with combined clinical factors and 19 proteins as features (R^2^_PDDS_=0.31, R^2^_PROMIS_ =0.35) again outperformed base models. The four LASSO models (PDDS, PROMIS; both binary and ordinal/continuous) with combined clinical and protein features shared 2 clinical features (disease subtype, disease duration) and 4 protein biomarkers (CDCP1, IL-12B, NEFL, PRTG).

**Conclusions:** Serum protein biomarker profiles improve the prediction of real-world MS disability status beyond clinical profile alone or clinical profile plus individual protein biomarker, reaching clinically actionable performance.

## Introduction

Multiple sclerosis (MS) is a chronic neurological disease that could cause progressive accumulation of neurological disability.^1,2^ People with MS (pwMS) exhibit individual variations in disease activity and trajectory.^3,4^ The available MS disease-modifying therapies (DMTs) pose soaring costs and exhibit variable real-world effectiveness in preventing inflammatory disease activity and delaying disability worsening.^5–7^ In the current practice, clinicians primarily rely on history, exams and neuroimaging to assess MS disease activity, disability progression, and treatment response. There is an unmet need to improve disease monitoring at the point of care to guide individualized management.

Blood biomarkers could potentially aid MS monitoring.^8–11^ Serum neurofilament light chain (sNEFL) and glial fibrillary acidic protein (sGFAP) are well-studied blood biomarkers of MS.^12–27^ NEFL and GFAP are neuron-specific and astrocyte-derived intermediate filament cytoskeletal proteins, respectively.^12,18^ Blood NEFL (*e*.*g*., sNEFL) has potential clinical applications in monitoring neuroaxonal damage associated with inflammatory disease activity (*i*.*e*., clinical and/or neuroimaging relapse), short-term disability worsening, and treatment response, though its utility to inform long-term disability remains unsettled. Blood GFAP (*e*.*g*., sGFAP) has potentially added utility in monitoring MS disability worsening, though it has limited ability to inform acute relapse. Despite these advances, few studies have examined the clinical utility of profiling multiple blood biomarkers beyond NEFL or GFAP in MS.

Leveraging the Proximity Extension Assay (PEA) methodology on the Olink™ platform,^28^ a prior study reported the analytical characterization of a custom serum-based proteomic multiplex immunoassay (PMI) panel (including sNEFL and sGFAP) associated with inflammatory MS disease activity and pertaining to key biological pathways in MS pathogenesis.^29^ Here, we deployed statistical learning methods to examine the association between serum biomarker profiles using the custom PMI panel and patient-reported disability in pwMS. Specifically, we hypothesized that serum protein profiles informative of inflammatory MS disease activity would also improve the prediction of real-world neurological disability when compared to clinical profiles or single serum protein.

## Methods

### Study Design and Cohorts

In this cross-sectional observational study (Figure 1A), we recruited participants from two centers: the University of Pittsburgh Medical Center (UPMC, n = 210) and the Rocky Mountain Multiple Sclerosis Clinic (RMMSC, n = 221) during 2017-2020. Inclusion criteria were adults 18 years or older with a neurologist-confirmed diagnosis of MS according to the 2017 McDonald criteria,^14^ clinically isolated syndrome (CIS), or radiologically isolated syndrome (RIS). We collected clinical and demographic data through review of electronic health records. Participants completed patient-reported outcomes (PROs) using either electronic or paper questionnaires. Participants donated venous blood samples during routine clinical appointments. Serum samples were isolated within 4 hours of phlebotomy and frozen at -80°C until proteomic profiling.

**Figure 1.**
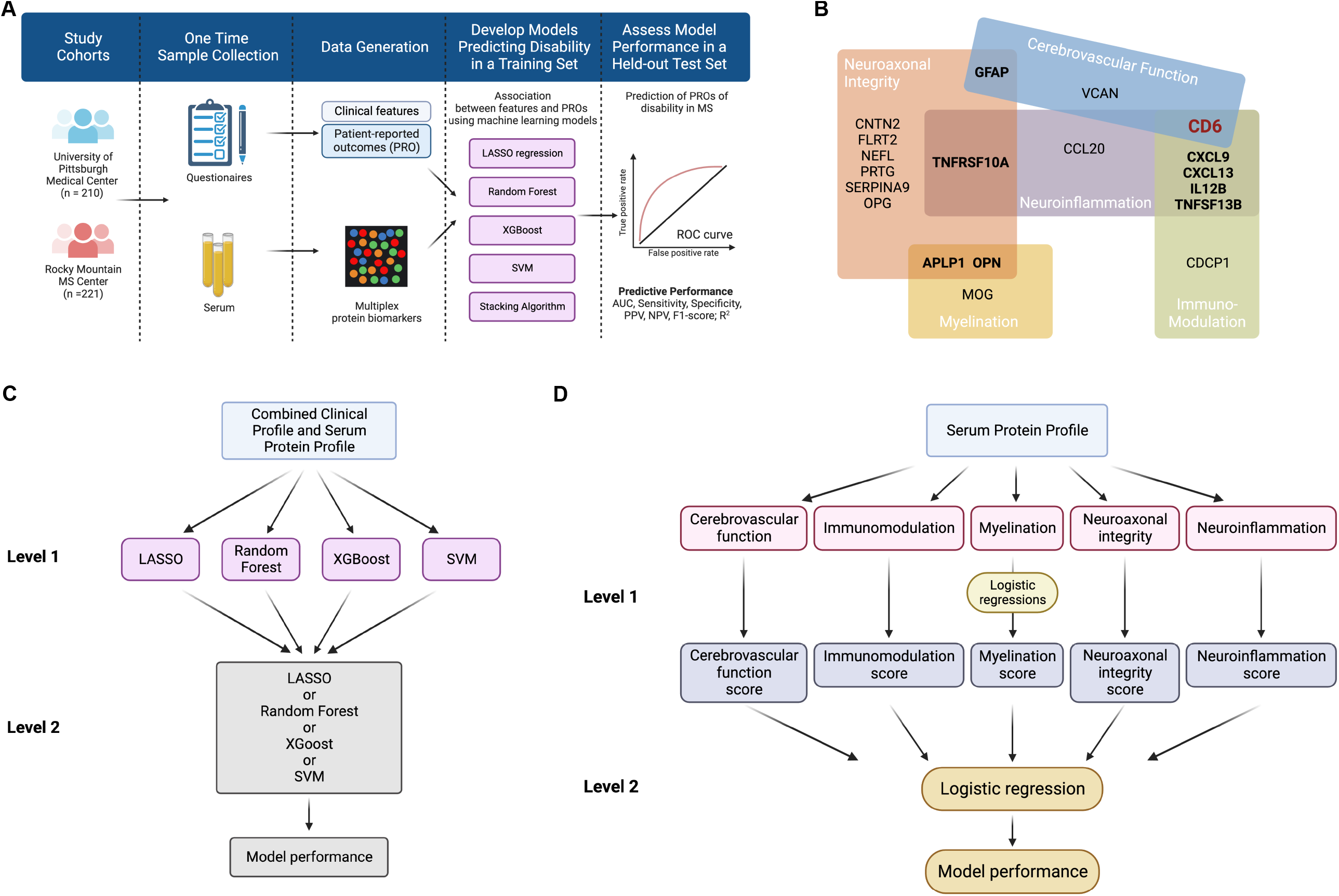
Overall study schematics. A. Study workflow. B. Functional pathways. The 19 protein biomarkers are grouped into 5 functional pathways: Cerebrovascular Function, Immunomodulation, Myelination, Neuroaxonal Integrity, and Neuroinflammation. C. Stacking ensemble learning. The Level 1 models include four different machine learning methods (LASSO, Random Forest, XGBoost and SVM) as ensemble members. The level 2 model then utilizes the predictions of the level 1 models to make the outcome prediction. D. Stacking classification algorithm using functional pathways as meta features in the predictive models. The level 1 logistic regression models use the biomarker concentrations as raw feature inputs to generate coefficients for biomarkers in each pathway (y = β_0_ + β_1_×biomaker_1_ + … + βn×biomaker_n_) and produce a probability score 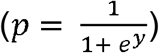 for the pathway The second level logistic regression uses the predicted probability scores of the functional pathways as meta-feature input.

### Ethics Approval

The institutional review boards of the University of Pittsburgh (STUDY19080007) and RMMSC (WCG20201562) approved the study protocols. All participants provided written informed consent.

### Serum Protein Biomarker Profile

Previously developed on the Olink™ platform using oligonucleotide-labeled antibodies and PEA methodology, a custom PMI panel comprising 19 proteins (see detailed protein names in eTable 1) measured the absolute concentration (pg/mL) of each serum protein.^13^ Briefly, in prior MS studies, a library of >1400 proteins was screened for association with standard MS disease activity endpoints, including clinically defined relapse versus remission, the presence (and count) versus absence of gadolinium-enhanced lesions on a matched magnetic resonance imaging (MRI), annualized relapse rate, and Expanded Disability Status Scale (EDSS).^30^ The custom panel included 19 proteins based on optimal performance for predicting these MS end points with a primary focus on inflammatory disease activity (*i*.*e*., clinical and/or radiological relapse). We assigned the 19 proteins to 5 biological pathways relevant to MS pathogenesis (Cerebrovascular Function, Immunomodulation, Myelination, Neuroaxonal Integrity, and Neuroinflammation) (Figure 1B).^31^ In this study, serum concentrations of the 19 proteins constituted the patient-level serum biomarker profile.

Serum samples were assayed in 5 batches. We did not adjust for batch in subsequent analyses because principal component analyses showed no significant batch effects (eFigure 1). We performed log transformation of protein concentrations to minimize outlier effects.

### Clinical Profile

We collected standard clinical and demographic features that may influence MS outcomes: age (at sample collection), sex (female versus male), race/ethnicity (non-Hispanic white versus otherwise), disease subtypes (RIS/CIS/relapse-remitting MS versus progressive MS), disease duration (years between MS diagnosis and sample collection), and DMT efficacy at PRO assessment (high-efficacy versus standard-efficacy versus no DMT) as well as the time interval between serum collection and the closest PRO assessment (when not on the same day). For DMT (used by the study populations), we categorized natalizumab, ocrelizumab, and rituximab as higher-efficacy, while dimethyl fumarate, fingolimod, glatiramer acetate, interferon beta and teriflunomide as standard-efficacy.

### Feature Sets

We created four feature sets: (1) clinical profile only, (2) serum biomarker profile containing 19 proteins, (3) clinical profile plus one protein (out of 19) at a time, and (4) combined clinical profile and serum biomarker profile (eTable 2). We compared the feature set combining the clinical profile and serum biomarker profile against other benchmark feature sets.

### Patient-Reported Outcomes (PROs) of Disability

We assessed real-world neurological and physical functions using two clinically relevant and interrelated PROs. First, the Patient Determined Disease Steps (PDDS) scale evaluates gait function, ranging from 0 to 8, with 0 indicating no gait impairment and 8 representing bed-bound status. PDDS approximates rater-assessed EDSS.^32^ Second, the National Institute of Health Patient-Reported Outcomes Measurement Information System (PROMIS) Physical Function (version 1.2) quantifies general physical function. Nationally validated, PROMIS is a computer-adaptive test to measure patient-reported health across a range of chronic diseases and demographics, including MS.^33,34^ PROMIS reports a T-score and standard deviation (SD) relative to the general US population, which has a mean T-score of 50 (SD=10). Higher PROMIS scores indicate better physical function or lower physical disability. While both are validated in pwMS, PDDS is MS-specific whereas PROMIS is a generalizable measure of real-world disability. Both study sites administered PDDS, while PROMIS was only available from UPMC. We included PROs surveyed on or after the blood sample collection day for analysis. We used PROs as both ordinal / continuous and binary variables. We dichotomized PDDS according to the requirement for full-time ambulatory assistance (≥4 versus <4) and PROMIS based on the disability severity (≥35 mild/moderate disability versus <35 severe disability).

### Training and Test Set

Given the clinical and demographic differences between the two cohorts (Table 1), we first split samples and data into 80:20 for a training and a held-out test set within each cohort. We then combined the training sets and held-out test sets from both cohorts into one training and one testing set for subsequent analyses using PDDS. For the subgroup analyses using PROMIS, we used the same 80:20 split for the UPMC cohort.

**Table 1.**
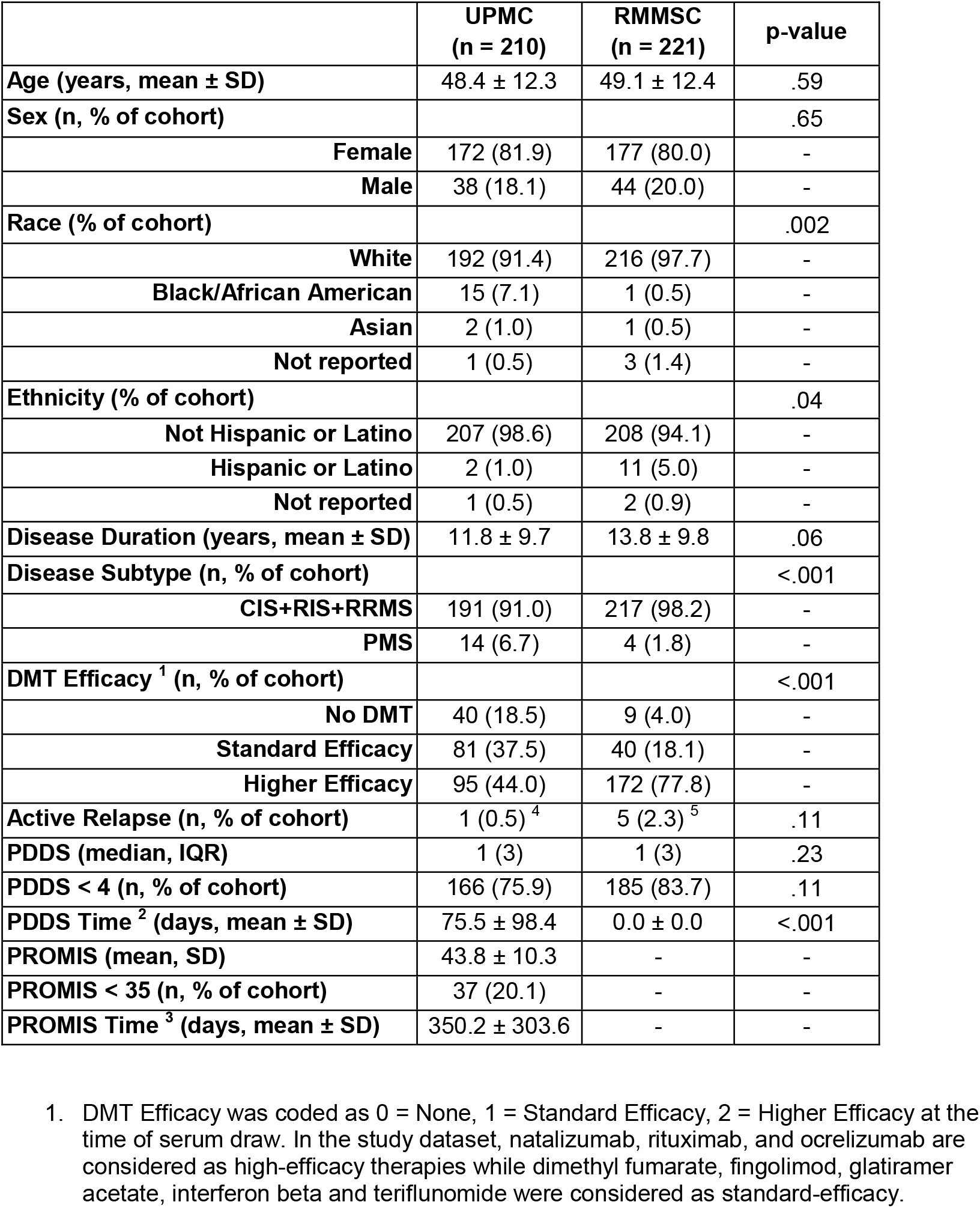
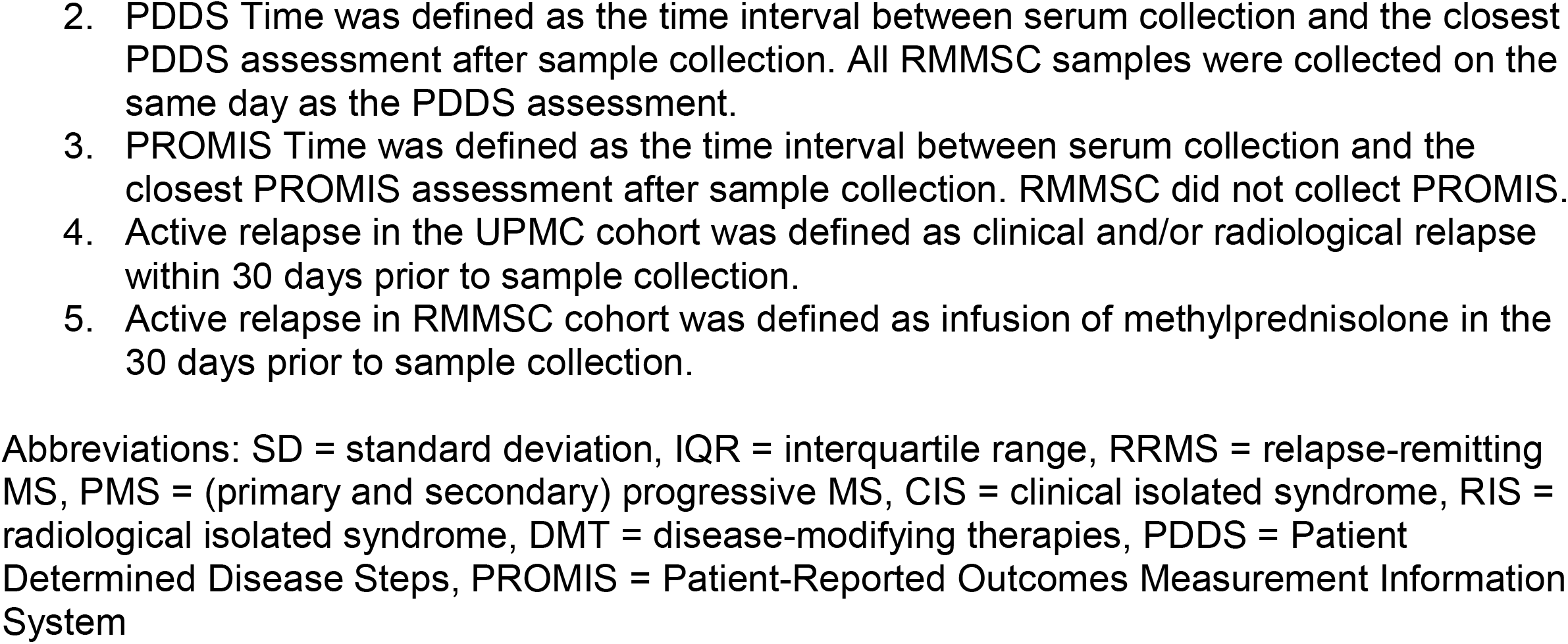
Patient Characteristics

|  | **UPMC**<br>**(n = 210)** | **RMMSC**<br>**(n = 221)** | **p-value** |
| --- | --- | --- | --- |
| **Age (years, mean ± SD)** | 48.4 ± 12.3 | 49.1 ± 12.4 | .59 |
| **Sex (n, % of cohort)** |  |  | .65 |
| **Female** | 172 (81.9) | 177 (80.0) | - |
| **Male** | 38 (18.1) | 44 (20.0) | - |
| **Race (% of cohort)** |  |  | .002 |
| **White** | 192 (91.4) | 216 (97.7) | - |
| **Black/African American** | 15 (7.1) | 1 (0.5) | - |
| **Asian** | 2 (1.0) | 1 (0.5) | - |
| **Not reported** | 1 (0.5) | 3 (1.4) | - |
| **Ethnicity (% of cohort)** |  |  | .04 |
| **Not Hispanic or Latino** | 207 (98.6) | 208 (94.1) | - |
| **Hispanic or Latino** | 2 (1.0) | 11 (5.0) | - |
| **Not reported** | 1 (0.5) | 2 (0.9) | - |
| **Disease Duration (years, mean ± SD)** | 11.8 ± 9.7 | 13.8 ± 9.8 | .06 |
| **Disease Subtype (n, % of cohort)** |  |  | <.001 |
| **CIS+RIS+RRMS** | 191 (91.0) | 217 (98.2) | - |
| **PMS** | 14 (6.7) | 4 (1.8) | - |
| **DMT Efficacy <sup>1</sup> (n, % of cohort)** |  |  | <.001 |
| **No DMT** | 40 (18.5) | 9 (4.0) | - |
| **Standard Efficacy** | 81 (37.5) | 40 (18.1) | - |
| **Higher Efficacy** | 95 (44.0) | 172 (77.8) | - |
| **Active Relapse (n, % of cohort)** | 1 (0.5) <sup>4</sup> | 5 (2.3) <sup>5</sup> | .11 |
| **PDDS (median, IQR)** | 1 (3) | 1 (3) | .23 |
| **PDDS < 4 (n, % of cohort)** | 166 (75.9) | 185 (83.7) | .11 |
| **PDDS Time <sup>2</sup> (days, mean ± SD)** | 75.5 ± 98.4 | 0.0 ± 0.0 | <.001 |
| **PROMIS (mean, SD)** | 43.8 ± 10.3 | - | - |
| **PROMIS < 35 (n, % of cohort)** | 37 (20.1) | - | - |
| **PROMIS Time <sup>3</sup> (days, mean ± SD)** | 350.2 ± 303.6 | - | - |

1. DMT Efficacy was coded as 0 = None, 1 = Standard Efficacy, 2 = Higher Efficacy at the time of serum draw. In the study dataset, natalizumab, rituximab, and ocrelizumab are considered as high-efficacy therapies while dimethyl fumarate, fingolimod, glatiramer acetate, interferon beta and teriflunomide were considered as standard-efficacy.
2. PDDS Time was defined as the time interval between serum collection and the closest PDDS assessment after sample collection. All RMMSC samples were collected on the same day as the PDDS assessment. 3. PROMIS Time was defined as the time interval between serum collection and the closest PROMIS assessment after sample collection. RMMSC did not collect PROMIS. 4. Active relapse in the UPMC cohort was defined as clinical and/or radiological relapse within 30 days prior to sample collection. 5. Active relapse in RMMSC cohort was defined as infusion of methylprednisolone in the 30 days prior to sample collection.
Abbreviations: SD = standard deviation, IQR = interquartile range, RRMS = relapse-remitting MS, PMS = (primary and secondary) progressive MS, CIS = clinical isolated syndrome, RIS = radiological isolated syndrome, DMT = disease-modifying therapies, PDDS = Patient Determined Disease Steps, PROMIS = Patient-Reported Outcomes Measurement Information System

### Machine Learning (ML) Methods

We deployed 6 different ML methods using multiple feature sets to predict PROs (eTable 2). ML methods included: (1) Least Absolute Shrinkage and Selection Operator (LASSO) regression, (2) Random Forest (RF), (3) XGBoost, (4) Support-Vector Machines (SVM), (5) stacking ensemble learning, and (6) stacking classification algorithm. LASSO performs penalized L1 regularization, which produces sparse models containing the minimal number of informative features.^35^ RF creates a collection of random uncorrelated decision trees to produce the best possible prediction.^36^ XGBoost optimally combines decision tree and linear regression under a Gradient Boosting framework, which efficiently decreases errors and effectively reduces irrelevant features.^37^ SVM performs supervised classifications to map features into discrete spaces to maximize the gap between data points in separate categories.^38^ Stacking ensemble learning combines the best predictions from two or more base ML methods (Figure 1C).^39^ For this study, stacking classification algorithm, which differs from stacking ensemble learning, enables the functional pathways as feature input in predictive models in two steps: (1) The first-level logistic regression models use biomarker concentrations as inputs to generate coefficients for the biomarkers in each given pathway and produce a probability score for the pathway; (2) The second-level logistic regression uses predicted probability scores of the functional pathways as meta features as model input (Figure 1B).^40^

To report model performance, we assessed the area under the receiver operating characteristic curve (AUC) with 95% confidence interval (CI) computed with 2,000 bootstrap replicates as well as sensitivity, specificity, positive predictive value (PPV) or precision, negative predictive value (NPV), and F1-score for *binary outcomes* as well as R^2^ with 95% CI for *continuous/ordinal outcomes*.

### Additional Statistical Analysis

We compared cohort characteristics using chi-square test for categorical data and t-test for continuous variables. We performed the Spearman correlation across all variables. For these tests, p-value <0.05 was deemed statistically significant. We performed all analyses using R, version 4.0.2.

## Results

### Patient Characteristics

The study included 431 participants with MS diagnosis (UPMC: n=210; RMMSC: n=221; Table 1). The two cohorts shared similar age, sex, and disease duration as well as the proportion with active relapse within 30 days prior to sample collection and the proportion with mild/moderate disability (PDDS<4). When compared to RMMSC, the UPMC cohort had a lower percentage of White (91.4% vs. 97.7%, p=.002), Hispanic/Latino (1.0% vs. 5.0%, p=.04) patients, higher percentage with progressive MS (6.7% vs. 1.8%, p<.001) and no DMT (18.5% vs. 4.0%, p<.001) or standard efficacy DMT (37.5% vs. 18.1%, p<.001) at sample collection. While all RMMSC participants completed PDDS assessment on the same day as sample collection, the time interval (mean±SD) between sample collection and the closest PDDS after sample collection was 75.5±98.4 days for UPMC participants. UPMC participants additionally completed PROMIS assessment, with a time interval of 350.2±303.6 days between sample collection and the closest PROMIS after sample collection.

### Feature Correlation

To assess the correlation structure among the model input features, we assessed pairwise correlations (eFigure 2). Among clinical features, disease duration and age showed the strongest correlation (r=0.57, p<.001). Among serum proteins, most significant correlations were positive, with MOG and APLP1 concentrations having the strongest correlation (r=0.67, p<.0001), while only GFAP and CD6 displayed inverse correlation (r=-0.39, p<.0001). When assessing serum proteins with clinical features, NEFL concentrations and age showed the strongest positive correlation (r=0.56, p<.001), whereas CXCL13 and DMT efficacy had the strongest inverse correlation (r=-0.26, p<.001).

### Best ML Model Performance for Informing Disability

The LASSO approach consistently outperformed the other ML approaches for predicting PROs of disability (Figure 2). Further, the combined clinical profile and serum biomarker profile (containing 19 proteins) consistently outperformed other benchmark feature sets.

**Figure 2.**
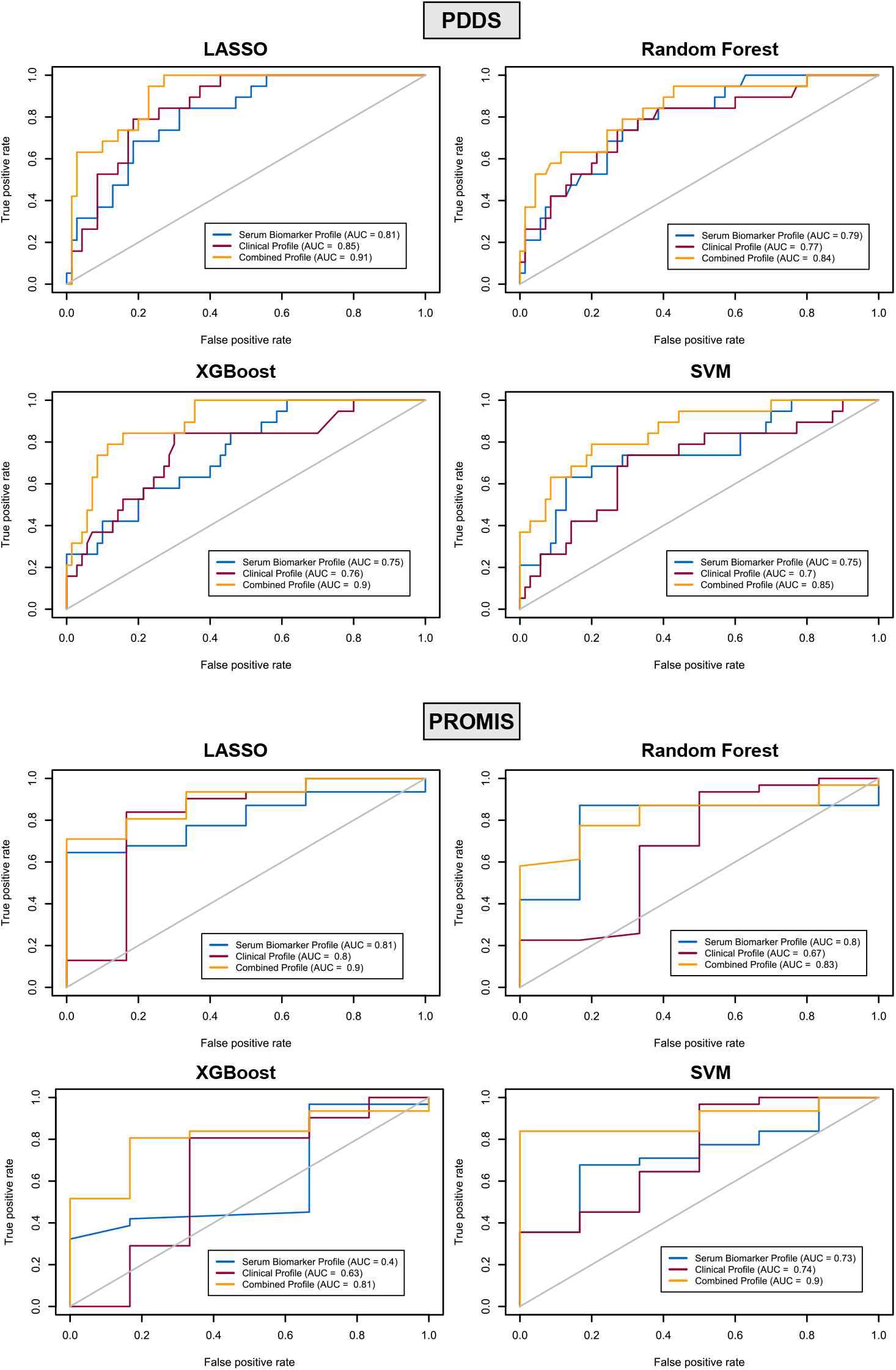
Receiver Operating Characteristic (ROC) plots of the different machine learning models for predicting patient-reported disability outcomes.

First, we assessed model predictive performance for MS-specific disability. When predicting severe versus mild/moderate MS disability (binary PDDS≥4 vs. <4), the LASSO model using the combined clinical profile and serum biomarker profile as feature input achieved the best AUC (0.91, 95%CI 0.85-0.97) as well as the best sensitivity (0.89), specificity (0.86), PPV (0.42), NPV (0.99), and F1-score (0.57) as compared to benchmark feature sets (i.e., clinical profile alone, serum biomarker profile alone; Table 2, eTable 3) or other ML models (eTable 6, eTable 7). Likewise, when predicting ordinal PDDS, the LASSO model using the combined feature set again achieved the best R^2^ (0.31, 95%CI 0.20-0.41) (Table 2, eTable 3, eTable 6). These two best performing models (LASSO using the combined feature set) to predict PDDS (as binary or ordinal outcome) selected slightly different sets of informative features (eTable 4 for feature coefficients). Collectively, both models shared the following input: 6 clinical features (age, sex, race/ethnicity, disease subtype, disease duration, DMT efficacy) and 9 proteins (CD6, CDCP1, CNTN2, IL12B, NEFL, PRTG, SERPINA9, TNFSF13B, VCAN) (eFigure 3).

**Table 2.** LASSO Model Predictive Performance and Selected Features for Patient-Reported Disability Outcomes

|  | PDDS Binary |  |  | PDDS Ordinal |  |  | PROMIS Binary |  |  | PROMIS Continuous |  |  |
| --- | --- | --- | --- | --- | --- | --- | --- | --- | --- | --- | --- | --- |
|  | Clinical Profile | Serum Protein Profile | Clinical Profile + Serum Protein Profile | Clinical Profile | Serum Protein Profile | Clinical Profile + Serum Protein Profile | Clinical Profile | Serum Protein Profile | Clinical Profile + Serum Protein Profile | Clinical Profile | Serum Protein Profile | Clinical Profile + Serum Protein Profile |
| **AUC (95% CI)** | 0.85<br>(0.77, 0.93) | 0.81<br>(0.71, 0.91) | 0.91<br>(0.85, 0.97) |  |  |  | 0.80<br>(0.53, 1.00) | 0.81<br>(0.66, 0.96) | 0.90<br>(0.78, 1.00) |  |  |  |
| **Sensitivity** | 0.60 | 0.71 | 0.89 |  |  |  | 0.89 | 0.89 | 0.93 |  |  |  |
| **Specificity** | 0.81 | 0.83 | 0.86 |  |  |  | 1.00<sup>x</sup> | 0.30 | 0.57 |  |  |  |
| **PPV** | 0.16 | 0.26 | 0.42 |  |  |  | 1.00<sup>x</sup> | 0.77 | 0.90 |  |  |  |
| **NPV** | 0.97 | 0.97 | 0.99 |  |  |  | 0.33 | 0.50 | 0.67 |  |  |  |
| **F1-score** | 0.25 | 0.38 | 0.57 |  |  |  | 0.94 | 0.83 | 0.92 |  |  |  |
| **R<sup>2</sup> (95% CI)** |  |  |  | 0.20<br>(0.07, .33) | 0.28<br>(0.16, 0.40) | 0.31<br>(0.20, 0.41) |  |  |  | 0.25<br>(0.06, 0.43) | 0.26<br>(0.16, 0.37) | 0.35<br>(0.29, 0.42) |
|  | **Clinical Profile Features**<sup>5,6</sup> |  |  |  |  |  |  |  |  |  |  |  |
| **Age** | Y | - | Y | Y | - | Y | Y | - | N | Y | - | Y |
| **Sex** | Y | - | Y | Y | - | Y | N | - | N | N | - | Y |
| **Race/Ethnicity** | Y | - | Y | Y | - | Y | N | - | N | N | - | N |
| **Disease Subtype<sup>1</sup>** | Y | - | Y | Y | - | Y | Y | - | Y | Y | - | Y |
| **Disease Duration** | Y | - | Y | Y | - | Y | Y | - | Y | N | - | Y |
| **DMT Efficacy<sup>2</sup>** | Y | - | Y | Y | - | Y | Y | - | Y | Y | - | N |
| **PDDS<sup>3</sup>/PROMIS<sup>4</sup> Time** | Y | - | Y | Y | - | N | N | - | N | Y | - | Y |
|  | **Serum Protein Profile Features**<sup>5,6</sup> |  |  |  |  |  |  |  |  |  |  |  |
| **APLP1** | - | N | N | - | Y | Y | - | Y | Y | - | Y | Y |
| **CCL20** | - | N | N | - | N | N | - | Y | N | - | Y | N |
| **CD6** | - | Y | Y | - | Y | Y | - | N | N | - | Y | Y |
| **CDCP1** | - | Y | Y | - | Y | Y | - | Y | Y | - | Y | Y |
| **CNTN2** | - | Y | Y | - | Y | Y | - | Y | Y | - | N | N |
| **CXCL13** | - | N | N | - | N | N | - | Y | N | - | N | N |
| **CXCL9** | - | Y | Y | - | Y | N | - | Y | N | - | N | N |
| **FLRT2** | - | N | N | - | N | N | - | Y | N | - | N | N |
| **GFAP** | - | Y | N | - | Y | Y | - | Y | Y | - | Y | Y |
| **IL12B** | - | Y | Y | - | Y | Y | - | Y | Y | - | Y | Y |
| **MOG** | - | Y | Y | - | N | N | - | N | N | - | N | N |
| **NEFL** | - | Y | Y | - | Y | Y | - | Y | Y | - | Y | Y |
| **OPG** | - | N | Y | - | N | N | - | N | N | - | Y | Y |
| **OPN** | - | N | N | - | N | N | - | Y | Y | - | Y | N |
| **PRTG** | - | Y | Y | - | Y | Y | - | Y | Y | - | Y | Y |
| **SERPINA9** | - | Y | Y | - | Y | Y | - | Y | Y | - | N | N |
| **TNFRSF10A** | - | Y | N | - | N | N | - | Y | N | - | Y | N |
| **TNFSF13B** | - | Y | Y | - | Y | Y | - | Y | N | - | Y | Y |
| **VCAN** | - | Y | Y | - | Y | Y | - | Y | N | - | Y | N |

1. Disease Subtype was coded as 0 = clinical isolated syndrome/radiological isolated syndrome/relapse-remitting MS, 1 = progressive MS at time of serum collection. 2. DMT Efficacy was coded as 0 = None, 1 = Standard Efficacy, 2 = Higher Efficacy of the DMT at the time of serum collection. In the study dataset, natalizumab, rituximab, and ocrelizumab are considered as high-efficacy therapies while dimethyl fumarate, fingolimod, glatiramer acetate, interferon beta and teriflunomide were considered as standard-efficacy. 3. PDDS Time was defined as the time interval between serum collection and the closest PDDS assessment after sample collection. 4. PROMIS Time was defined as the time interval between serum collection and the closest PROMIS assessment after sample collection. 5. Y indicates features selected by LASSO models. 6. N indicates features NOT selected by LASSO models.
<sup>x</sup> The performance of the LASSO model for the binary PROMIS (specificity and PPV) might be influenced by the small sample size of the testing set (n = 20).
Abbreviations: AUC = area under the ROC curve, 95% CI = 95% confidence interval, PPV = positive predictive value, NPV = negative predictive value, PDDS = Patient Determined Disease Steps, PROMIS = Patient-Reported Outcomes Measurement Information System

To further explore the contribution of each serum protein to model performance, we systematically assessed LASSO models using clinical profile plus *individual* protein (one at a time rather than the serum biomarker profile comprising all 19 proteins) as feature input for predicting severe disability (binary PDDS≥4 vs. <4). The addition of a single protein did not significantly improve LASSO model performance beyond the clinical profile alone (AUC 0.85, 95%CI 0.77-0.93) (eTable 3). Notably, LASSO models using clinical profile plus individual protein as feature input did not select five proteins as the final features, including NEFL and GFAP. When forcing either NEFL or GFAP (as individual protein) into the respective LASSO models, none of the model performance metrics improved beyond the base model using clinical profile alone (eTable 3).

Next, we assessed model predictive performance for general physical disability using PROMIS in subgroup analyses (UPMC cohort, n=210). Again, LASSO models using the combined clinical profile and serum biomarker profile as the feature input showed the best performance (binary PROMIS <35 vs. ≥35: AUC 0.90, 95%CI 0.78-1.00; continuous PROMIS: R^2^ 0.35, 95%CI 0.29-0.42) as compared to other benchmark feature sets or other ML models (Table 2, eTable 6). These two models (LASSO using the combined feature set) to predict PROMIS (as binary or continuous outcome) also selected slightly different informative features (eTable 5 for feature coefficients), and collectively shared 2 clinical features (disease duration, disease subtype) and 6 proteins (APLP1, CDCP1, GFAP, IL12B, NEFL, PRTG) (eFigure 3).

When assessing the overlap of the features selected by the four LASSO models (using the combined feature set to predict PDDS and PROMIS, binary and ordinal/continuous), 2 clinical features (disease subtype, disease duration) and 4 proteins (CDCP1, IL-12B, NEFL, PRTG) persisted (eFigure 3).

### Other Machine Learning and Ensemble Approaches for Predicting Disability

When compared to LASSO, the other ML approaches (RF, XGBoost, SVM) consistently performed worse for predicting both severe MS disability and severe general physical disability across all performance metrics (binary PDDS, binary PROMIS: Figure 2, eTable 6). Like LASSO models, the combined feature set as input consistently outperformed benchmark feature sets when using RF, XGBoost or SVM. While XGBoost (using the combined feature set) achieved similar AUCs as LASSO, it had markedly worse sensitivity. SVM (using the combined feature set) achieved similar sensitivity, specificity and NPV as LASSO, but achieved worse AUC.

We further tested whether a stacking ensemble approach using the combined feature sets could further improve the predictive performance over the individual ML models (LASSO, RF, XGBoost, or SVM). The stacking ensemble approach did not perform better than the best individual ML approach (LASSO) for predicting MS-specific or general disability (binary PDDS, binary PROMIS: Figure 2, eTable 7). The lack of significant improvement could be due to the already robust performance of the individual ML methods.

### Predictive Performance Using Functional Pathways

Given that the 19 proteins in the serum protein profile can be organized into 5 functional pathways, we examined the predictive performance of the 5 functional pathways as meta-feature input in predicting severe MS disability (binary PDDS) using the stacking classification algorithm. The stacking classification algorithm using clinical profile and 5 functional pathways as feature input showed better model performance (AUC 0.86, 95%CI 0.74-0.86; sensitivity 0.95; specificity 0.62, PPV 0.44, NPV 0.98, F1-score 0.61) than the model using 5 functional pathways alone (AUC 0.77, 95%CI 0.65-0.77; sensitivity 0.81; specificity 0.61). When compared with LASSO model using combined clinical and protein profiles (AUC 0.91, 95%CI 0.85-0.97; sensitivity 0.89; specificity 0.62, PPV 0.42, NPV 0.99, F1-score 0.57), the stacking classification algorithm combining clinical profile and 5 functional pathways as feature input achieved worse AUC but better sensitivity and comparable specificity, PPV, NPV and F1-score (Table 2-3). The immunomodulation (p=.02) and the neuroaxonal integrity (p<.001) pathways significantly contributed to the predictive performance of the stacking classification algorithm using 5 functional pathways alone, while the neuroaxonal integrity (p<.001) pathway remained significant in the model comprising the combined clinical profile and the 5 functional pathways. Notably, the four protein biomarkers selected by 4 LASSO models using combined clinical and protein profiles (Table 2, eFigure 3) are involved in either neuroaxonal integrity pathway (NEFL and PRTG) or immunomodulation pathway (CDCP1 and IL-12B). The neuroaxonal integrity pathway also includes 4 proteins (APLP1, CNTN2, SERPINA9, TNFSF13B) shared by two or more best performing LASSO models in predicting PDDS or PROMIS (eFigure 3).

**Table 3.** Model Performance of the Stacking Classification Algorithm and the Significant Functional Pathways for Predicting Patient Determined Disease Steps

|  | Functional Pathways |  | Functional Pathways + Clinical Profile |  |
| --- | --- | --- | --- | --- |
| AUC (95% CI) | 0.77 (0.65, 0.77) |  | 0.86 (0.74, 0.86) |  |
| Sensitivity | 0.81 |  | 0.95 |  |
| Specificity | 0.61 |  | 0.62 |  |
| PPV | 0.40 |  | 0.44 |  |
| NPV | 0.91 |  | 0.98 |  |
| F1-score | 0.53 |  | 0.61 |  |
|  | Coefficient | P-value | Coefficient | P-value |
|  | Clinical Features <sup>1</sup> |  |  |  |
| Age | - | - | 0.040 | 0.034 |
| Sex | - | - | -0.474 | 0.254 |
| Race / Ethnicity | - | - | -0.474 | 0.254 |
| Disease Subtype | - | - | 0.689 | 0.051 |
| Disease Duration | - | - | 0.039 | 0.026 |
| DMT Efficacy | - | - | 0.404 | 0.138 |
| PDDS Time | - | - | 0.005 | 0.004 |
|  | Functional Pathways <sup>2</sup> |  |  |  |
| Neuroaxonal Integrity | 0.903 | < .001 | 0.653 | 0.006 |
| Immunomodulation | 0.798 | 0.024 | 0.459 | 0.251 |
| Myelination | -0.996 | 0.392 | -1.566 | 0.218 |
| Neuroinflammation | 0.225 | 0.595 | 0.304 | 0.521 |
| Cerebrovascular Function | 1.256 | 0.766 | 1.157 | 0.199 |

1. Please refer to Methods and Table 1 for description of the clinical features. 2. Please refer to Figure 1C and eTable 1 for the proteins of each functional pathway.
Abbreviations: AUC = area under the ROC curve, 95% CI = 95% confidence interval, PPV = positive predictive value, NPV = negative predictive value

## Discussion

The key study finding is that the addition of serum biomarker profiles comprising multiple proteins associated with MS inflammatory disease activity improved the machine learning model performance of predicting real-world disability status beyond clinical profile alone, reaching clinically actionable accuracy (as well as other performance metrics). Importantly, serum biomarker profiles consistently outperformed individual serum proteins such as NEFL or GFAP as model feature input. Proteins involved in neuroaxonal integrity significantly contributed to the predictive performance of serum biomarker profile when combined with clinical profiles. LASSO consistently outperformed other machine learning approaches, including RF, XGBoost or SVM.

Our study has several strengths. First, we leveraged data from two independent cohorts with clinically distinct characteristics. Specifically, we reported the machine learning model performance using the held-out test sets. Thus, the study findings have potential generalizability. Second, we employed two PROs of disability to represent real-world evidence of functional status. Both PDDS and PROMIS are well validated in pwMS.^32–34^ Few prior studies examined the clinical utility of blood biomarkers in predicting PROs in MS, and none focused on patient-reported disability status. Third, we applied multiple machine learning approaches that all confirmed the added utility of serum biomarker profiles as feature input in predicting disability in pwMS beyond clinical profile alone. Finally, this is the first study to our knowledge that demonstrates the potential clinical application of serum biomarker profile comprising multiple proteins in predicting real-world MS disability status, outperforming individual protein biomarkers. Prior studies showed that individual blood protein biomarker such as sNEFL or sGFAP alone was insufficient to accurately predict MS disability progression or treatment response.^14,16,18,41^ In our study, combined feature sets comprising key clinical features and multiple protein biomarkers consistently outperformed not only clinical profile alone but importantly also clinical profile plus individual protein (including sNEFL or sGFAP) in predicting patient-reported disability in pwMS. These findings suggest that serum biomarker profiles comprising multiple proteins better capture the complex disease states of pwMS (*i*.*e*., disability) and may have clinical application in real-world monitoring of MS.

Notably, the protein biomarkers (CDCP1, IL-12B, NEFL and PRTG) selected by the four best performing LASSO models (for predicting PDDS or PROMIS, either as binary or ordinal/continuous outcomes) are involved in the two functional pathways (immunomodulation, neuroaxonal integrity) that significantly contributed to the predictive performance of stacking classification algorithm. In particular, NEFL and PRTG (pertinent to neuroaxonal integrity) significantly contributed to the performance of the stacking classification algorithm containing the five functional pathways in conjunction with clinical profile. Further, four other proteins (APLP1, CNTN2, GFAP and SERPINA9) selected by two or more best performing LASSO models (for predicting PDDS or PROMIS) are also involved in the same neuroaxonal integrity functional pathway. In prior studies, these four proteins are associated with MS inflammatory disease activity.^30^

Further, the three proteins (APLP1, CNTN2, GFAP) in addition to the best known NEFL play roles in MS pathogenesis, including demyelination and remyelination,^42,43^ grey matter pathology,^44^ and T cell dysregulation.^45,46^ Given the complex pathogenesis of MS, our findings suggested that a serum biomarker profile encompassing proteins involved in different MS pathogenic pathways is better than a single protein biomarker in informing real-world MS disability status.

Our study also has limitations. First, the current cross-sectional study design does not allow testing predictions of disability progression. Long-term follow-up study is under way to address this in future studies. Second, disability status based on PROs might contain ascertainment or other bias. On the other hand, both PDDS and PROMIS are well-validated against rater-assessed exams and provide critical real-world evidence of neurological function.^32–34^ Third, the limited racial and ethnic diversity of the two study cohorts reduced the generalizability beyond non-Hispanic white participants.

## Conclusion

Serum protein biomarker profiles based on proteomic multiplex immunoassay improve beyond clinical profile alone or clinical profile plus individual protein biomarker in informing real-world MS disability status, reaching clinically actionable performance. Future studies that include long-term follow-up from baseline clinical profile and serum biomarker profile ascertainment, incorporate objective functional testing in conjunction with PROs, and recruit higher proportions of participants from more diverse racial and ethnic backgrounds would further establish the clinical utility of this integrated approach in monitoring individual MS disease trajectories, particularly in predicting relapse-free disability progression.

## Data Availability

Code for analysis and figures is available at < https://github.com/xialab2016/MSbiomarker.git>. De-identified data are available upon request to the corresponding author and with permission from the participating institution.

## Acknowledgment

The authors thank all research participants and the clinicians from UPMC and RMMSC.

