## Supplemental Figures and Tables for "Association between Serum Biomarker Profile and Real-World Evidence of Disability in Multiple Sclerosis"

**SUPPLEMENTARY FIGURES AND TABLES**

**eFigure 1. Principal component analysis of serum sampling** showed no significant batch effects.

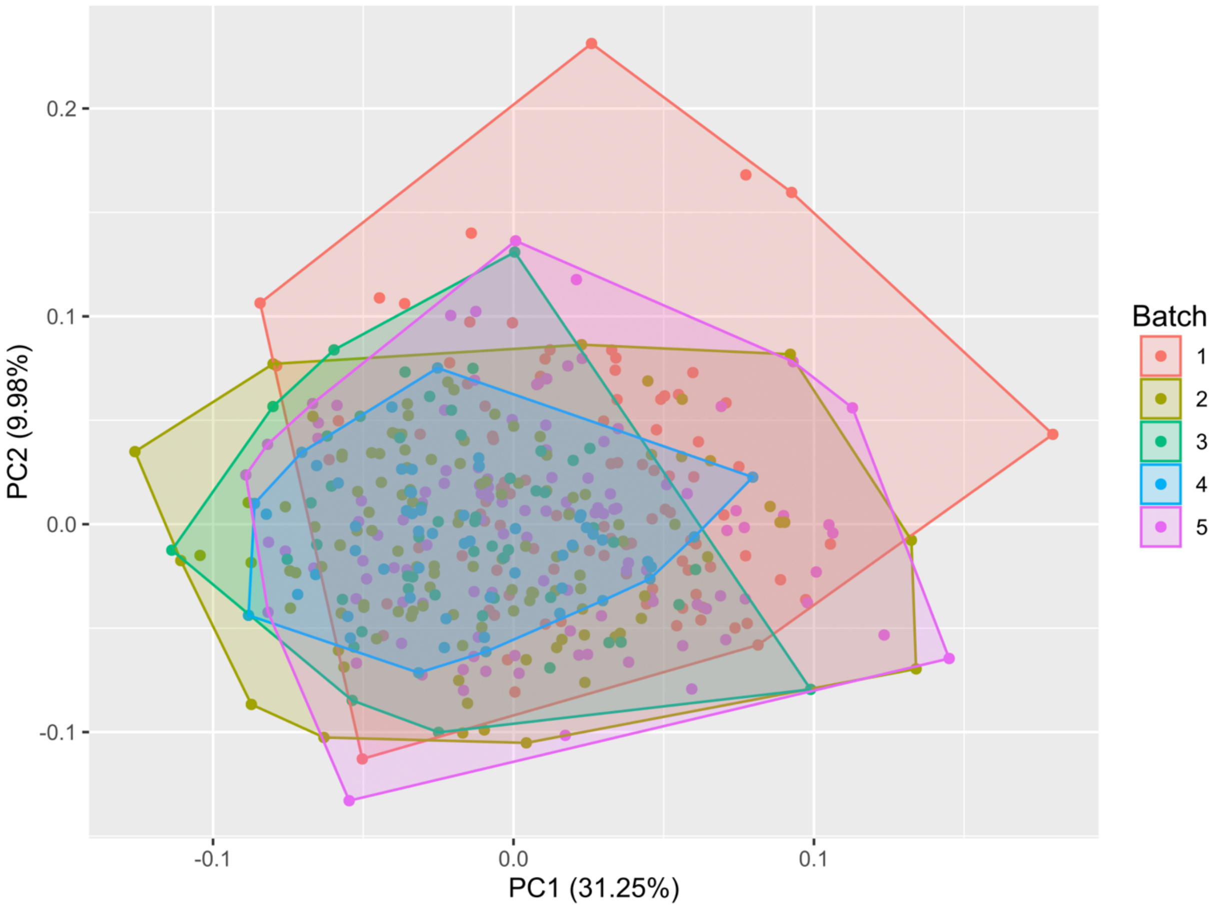

**eFigure 2. Correlation structure of features.**

Each square shows the pairwise Spearman correlation coefficient. Presence of the color indicates nominal significance (p<.05) with blue indicating positive and red indicating inverse correlation.

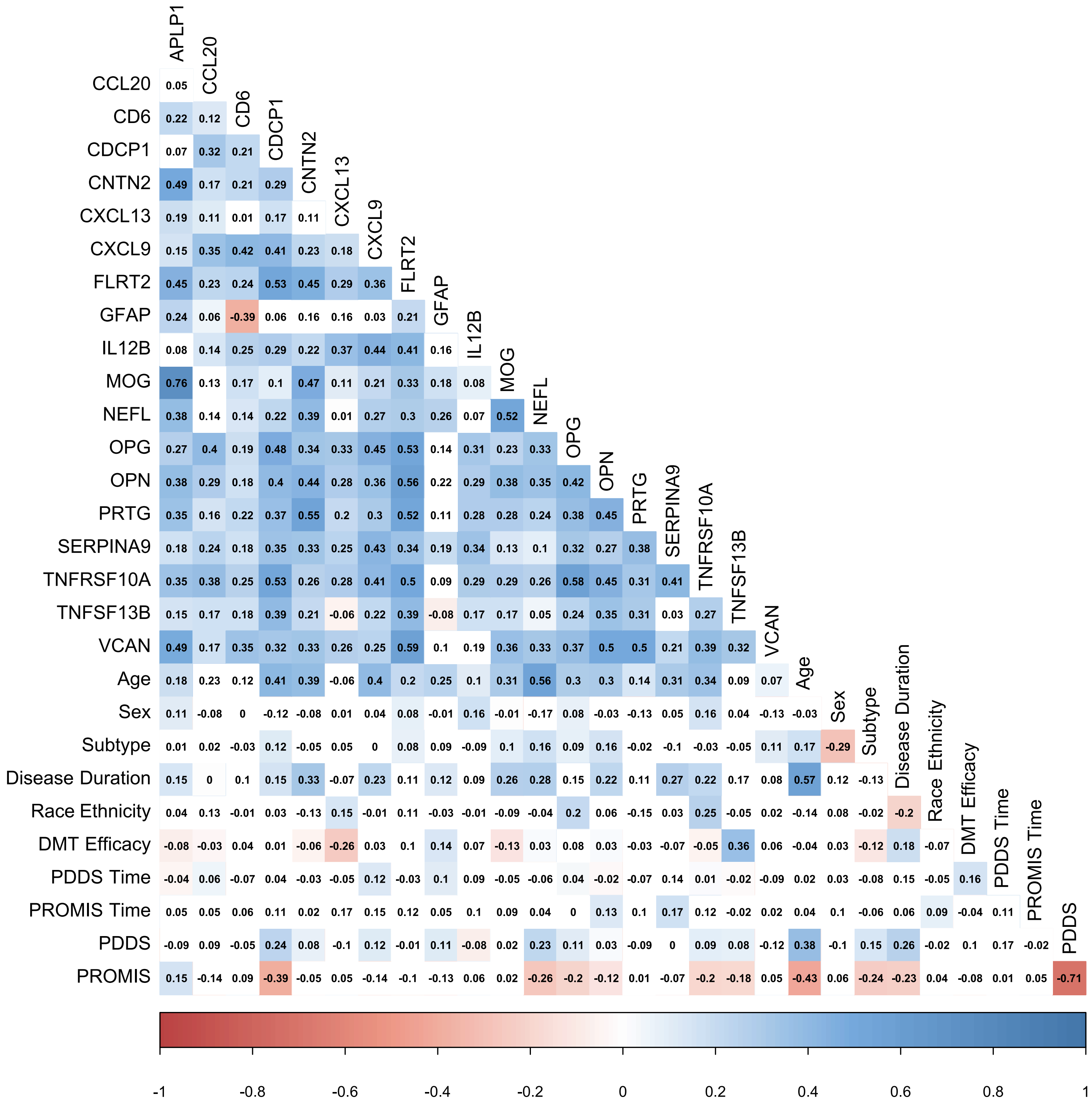

**eFigure 3. Features shared by different LASSO models.**
Features in **boldface**: shared by all four LASSO models; Features in *italics*: shared by three LASSO models; Features with underscore: shared by two LASSO models.

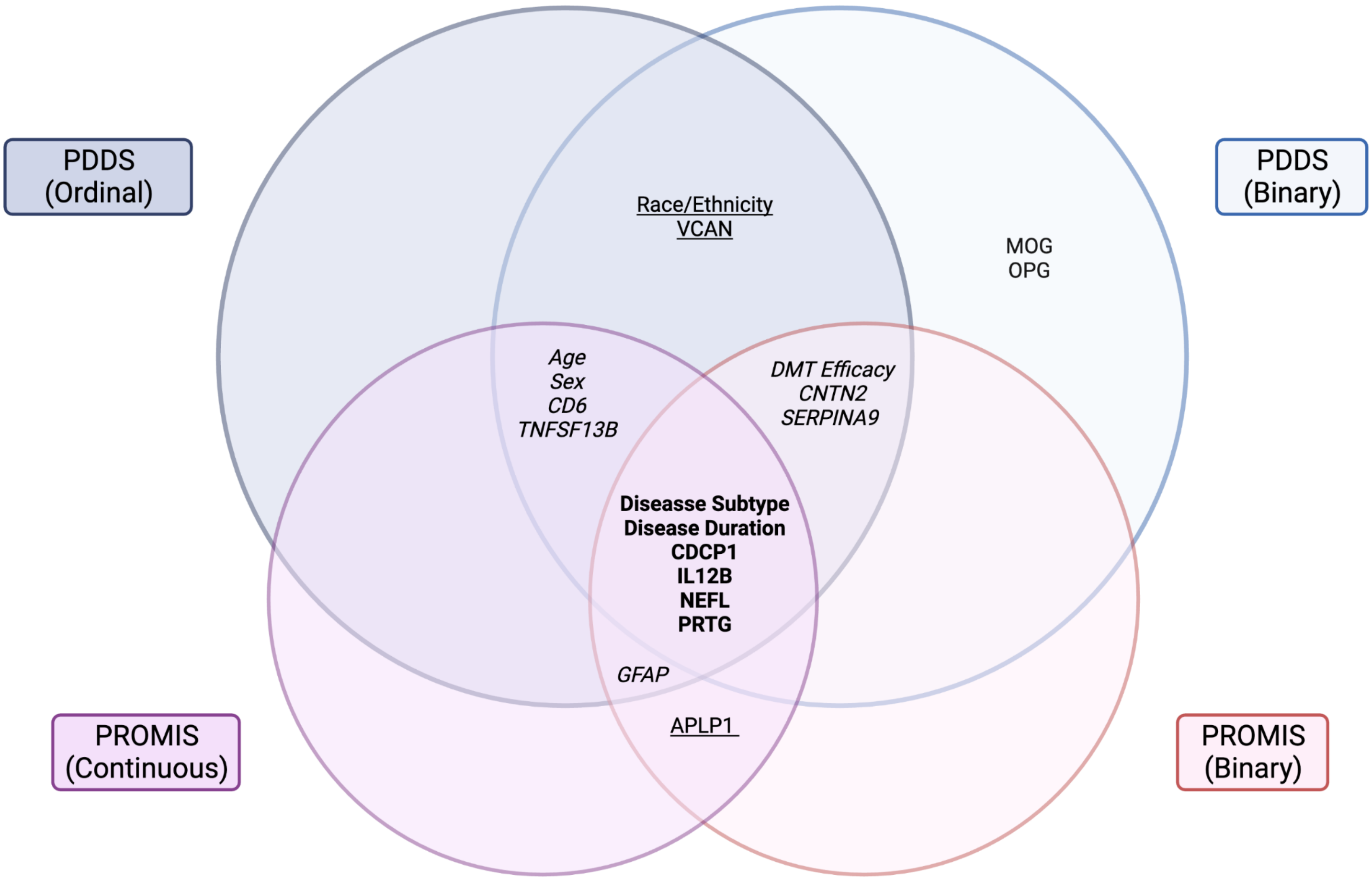

**eTable 1.** The 19 protein biomarkers and their associated functional pathways.

| Abbreviation | Protein Name | Functional Pathway(s) [1] |
|---|---|---|
| **APLP1** | Amyloid-like protein 1 | Myelination, Neuroaxonal Integrity |
| **CCL20** | Chemokine (C-C motif) ligand 20, | Neuroinflammation |
| **CD6** | Cluster of differentiation 6 | Cerebrovascular Function, Immunomodulation, Neuroaxonal Integrity |
| **CDCP1** | CUB domain-containing protein 1 | Immunomodulation |
| **CNTN2** | Contactin-2 | Neuroaxonal Integrity |
| **CXCL13** | Chemokine (C-X-C motif) ligand 13, | Immunomodulation, Neuroinflammation |
| **CXCL9** | Chemokine (C-X-C motif) ligand 9 | Immunomodulation, Neuroinflammation |
| **FLRT2** | Fibronectin leucine-rich repeat transmembrane protein 2 | Neuroaxonal Integrity |
| **GFAP** | Glial fibrillary acidic protein | Cerebrovascular Function, Neuroaxonal Integrity |
| **IL12B** | Interleukin-12 subunit beta | Immunomodulation, Neuroinflammation |
| **MOG** | Myelin oligodendrocyte glycoprotein | Myelination |
| **NEFL** | Neurofilament light chain | Neuroaxonal Integrity |
| **OPG** | Osteoprotegerin | Neuroaxonal Integrity, Neuroinflammation |
| **OPN** | Osteopontin | Myelination, Neuroaxonal Integrity |
| **PRTG** | Protogenin | Neuroaxonal Integrity |
| **SERPINA9** | Serpin family A member 9 | Neuroaxonal Integrity |
| **TNFSF10A** | Tumor necrosis factor ligand superfamily member 10 | Neuroaxonal Integrity, Neuroinflammation |
| **TNFSF13B** | Tumor necrosis factor ligand superfamily member 13B | Immunomodulation, Neuroinflammation |
| **VCAN** | Versican core protein | Cerebrovascular Function |

1. Please refer to the Methods section in the main text and main Figure 1B for functional pathways. A given protein may belong to more than one pathway.

**eTable 2.** Feature sets for the different machine learning models.

| Machine Learning Models [1] | Feature Sets | | | | |
|---|---|---|---|---|---|
| | **Clinical Profile** | **Single Serum Protein** | **Serum Protein Profile** | **Functional Pathways** | **Combined Profile** |
| **LASSO** | X | X | X | | X |
| **Random Forest** | X | | X | | X |
| **XGBoost** | X | | X | | X |
| **Support Vector Machine** | X | | X | | X |
| **Stacking Ensemble Learning** | | | | | X |
| **Stacking Classification** | | | | X | X |

1. Please refer to the Methods section in the main text for description of the different machine learning models.

**eTable 3.** Predictive performance and feature coefficient of benchmark LASSO models using combined clinical profile and a single protein marker (at a time) as feature input.

### eTable 3a

| | | | Coefficient = 0 [1] | | | | | | Forced [2] | |
|---|---|---|---|---|---|---|---|---|---|---|
| | Clinical Profile | | CCL20 | CXCL13 | OPN | NEFL | GFAP | | NEFL | GFAP |
| AUC (95% CI) | 0.85 (0.77, 0.93) | | 0.86 (0.78, 0.94) | 0.85 (0.77, 0.93) | 0.86 (0.78, 0.94) | 0.86 (0.78, 0.94) | 0.85 (0.77, 0.93) | | 0.85 (0.77, 0.93) | 0.85 (0.77, 0.93) |
| Sensitivity | 0.60 | | 0.75 | 0.75 | 0.75 | 0.75 | 0.75 | | 0.60 | 0.60 |
| Specificity | 0.81 | | 0.81 | 0.81 | 0.81 | 0.81 | 0.81 | | 0.81 | 0.81 |
| PPV | 0.16 | | 0.16 | 0.16 | 0.16 | 0.16 | 0.16 | | 0.16 | 0.16 |
| NPV | 0.97 | | 0.99 | 0.99 | 0.99 | 0.99 | 0.99 | | 0.97 | 0.97 |
| F1-score | 0.25 | | 0.26 | 0.26 | 0.26 | 0.26 | 0.26 | | 0.25 | 0.25 |
| Protein | NA | | NS | NS | NS | NS | NS | | -0.00036 | 0.00044 |
| Age | 0.00944 | | 0.00902 | 0.00926 | 0.00902 | 0.01000 | 0.01000 | | 0.00948 | 0.00944 |
| Sex | -0.07995 | | -0.06443 | -0.07331 | -0.06443 | -0.07000 | -0.07000 | | -0.08120 | -0.08025 |
| Race / Ethnicity | 0.08149 | | 0.07336 | 0.07801 | 0.07336 | 0.07000 | 0.08000 | | 0.08214 | 0.08159 |
| Disease Subtype | 0.00001 | | 0.00001 | 0.00001 | 0.00001 | NS | NS | | 0.00001 | 0.00001 |
| Disease Duration | 0.04749 | | 0.01828 | 0.03499 | 0.01828 | 0.02000 | 0.03000 | | 0.04984 | 0.04791 |
| DMT Efficacy | 0.05007 | | 0.03867 | 0.04519 | 0.03867 | 0.04000 | 0.04000 | | 0.05095 | 0.05015 |
| PDDS Time | 0.00033 | | 0.00026 | 0.00030 | 0.00026 | NS | NS | | 0.00034 | 0.00033 |

### eTable 3b

| | Coefficient ≠ 0 [3] | | | | | | |
|---|---|---|---|---|---|---|---|
| | **APLP1** | **CD6** | **CDCP1** | **CNTN2** | **CXCL9** | **FLRT2** | **IL12B** |
| **AUC (95% CI)** | 0.85 (0.78, 0.93) | 0.86 (0.78, 0.94) | 0.85 (0.76, 0.93) | 0.85 (0.76, 0.93) | 0.84 (0.76, 0.92) | 0.85 (0.77, 0.93) | 0.85 (0.76, 0.93) |
| **Sensitivity** | 0.80 | 0.75 | 0.67 | 0.75 | 0.62 | 0.67 | 0.67 |
| **Specificity** | 0.82 | 0.81 | 0.82 | 0.81 | 0.83 | 0.82 | 0.84 |
| **PPV** | 0.21 | 0.16 | 0.21 | 0.16 | 0.26 | 0.21 | 0.32 |
| **NPV** | 0.99 | 0.99 | 0.97 | 0.99 | 0.96 | 0.97 | 0.96 |
| **F1-score** | 0.33 | 0.26 | 0.32 | 0.26 | 0.37 | 0.32 | 0.43 |
| **Protein** | -0.06045 | -0.07983 | 0.03346 | 0.02614 | 0.02399 | -0.03496 | -0.04675 |
| **Age** | 0.00969 | 0.00931 | 0.00886 | 0.00926 | 0.00926 | 0.00945 | 0.00986 |
| **Sex** | -0.07568 | -0.09548 | -0.07827 | -0.08063 | -0.08832 | -0.07359 | -0.07939 |
| **Race/Ethnicity** | 0.08656 | 0.08467 | 0.07951 | 0.08049 | 0.08514 | 0.08172 | 0.08716 |
| **Disease Subtype** | 0.00001 | 0.00002 | 0.00001 | 0.00001 | 0.00001 | 0.00001 | 0.00001 |
| **Disease Duration** | 0.06479 | 0.06653 | 0.04719 | 0.04817 | 0.06264 | 0.04335 | 0.06015 |
| **DMT Efficacy** | 0.05184 | 0.05033 | 0.05228 | 0.05210 | 0.05675 | 0.04710 | 0.05904 |
| **PDDS Time** | 0.00036 | 0.00035 | 0.00033 | 0.00034 | 0.00032 | 0.00032 | 0.00039 |
| | **Coefficient ≠ 0 [3]** | | | | | | |
| | **MOG** | **OPG** | **PRTG** | **SERPINA9** | **TNFRSF10A** | **TNFSF13B** | **VCAN** |
| **AUC (95% CI)** | 0.85 (0.77, 0.93) | 0.85 (0.77, 0.93) | 0.86 (0.77, 0.94) | 0.85 (0.76, 0.93) | 0.85 (0.77, 0.93) | 0.85 (0.76, 0.93) | 0.87 (0.80, 0.95) |
| **Sensitivity** | 0.57 | 0.80 | 0.50 | 0.71 | 0.75 | 0.71 | 0.71 |
| **Specificity** | 0.82 | 0.82 | 0.81 | 0.83 | 0.81 | 0.83 | 0.83 |
| **PPV** | 0.21 | 0.21 | 0.16 | 0.26 | 0.16 | 0.26 | 0.26 |
| **NPV** | 0.96 | 0.99 | 0.96 | 0.97 | 0.99 | 0.97 | 0.97 |
| **F1-score** | 0.31 | 0.33 | 0.24 | 0.38 | 0.26 | 0.38 | 0.38 |
| **Protein** | -0.12751 | 0.01514 | -0.08785 | -0.02451 | -0.05513 | 0.07100 | -0.15700 |
| **Age** | 0.01062 | 0.00927 | 0.00958 | 0.00962 | 0.01003 | 0.00966 | 0.00962 |
| **Sex** | -0.07055 | -0.07843 | -0.08707 | -0.07804 | -0.07697 | -0.08279 | -0.08469 |
| **Race/Ethnicity** | 0.08758 | 0.07893 | 0.08351 | 0.08196 | 0.08552 | 0.07128 | 0.08795 |
| **Disease Subtype** | 0.00001 | 0.00001 | 0.00001 | 0.00001 | 0.00001 | 0.00001 | 0.00001 |
| **Disease Duration** | 0.05503 | 0.03817 | 0.04663 | 0.05080 | 0.06932 | 0.04979 | 0.04337 |
| **DMT Efficacy** | 0.04890 | 0.04771 | 0.04994 | 0.04812 | 0.05019 | 0.04169 | 0.04720 |
| **PDDS Time** | 0.00035 | 0.00031 | 0.00034 | 0.00034 | 0.00037 | 0.00032 | 0.00034 |

1. Protein biomarkers with coefficients of 0 were not selected by the individual models.
2. LASSO models in which either NEFL or GAFP were forced into the model.
3. Protein biomarkers with non-zero coefficients were selected by the individual models.

Abbreviations: AUC = area under the ROC curve, 95% CI = 95% confidence interval, PPV = positive predictive value, NPV = negative predictive value

**eTable 4.** Feature coefficient of LASSO models predicting PDDS.

| | PDDS Binary | | | PDDS Ordinal | | |
|---|---|---|---|---|---|---|
| | Clinical Profile | Serum Protein Profile | Clinical Profile + Serum Protein Profile | Clinical Profile | Serum Protein Profile | Clinical Profile + Serum Protein Profile |
| | Clinical Features [1] | | | | | |
| **Age** | 0.00944 | - | 0.00824 | 0.03861 | - | 0.02797 |
| **Sex** | -0.07995 | - | -0.05932 | -0.30695 | - | -0.22614 |
| **Subtype** | 0.08149 | - | 0.06346 | 0.22352 | - | 0.14558 |
| **Disease Duration** | 0.00001 | - | 0.00001 | 0.00006 | - | 0.00006 |
| **Race/Ethnicity** | 0.04749 | - | 0.00880 | 0.23536 | - | 0.08613 |
| **DMT Efficacy** | 0.05007 | - | 0.02673 | 0.14893 | - | 0.05732 |
| **PDDS Time** | 0.00033 | - | 0.00016 | NS | - | NS |
| | Protein Features [2] | | | | | |
| **APLP1** | - | NS | NS | - | -0.30884 | -0.14282 |
| **CCL20** | - | NS | NS | - | NS | NS |
| **CD6** | - | -0.09263 | -0.06976 | - | -0.16004 | -0.12296 |
| **CDCP1** | - | 0.09062 | 0.02342 | - | 0.69860 | 0.46535 |
| **CNTN2** | - | 0.09778 | 0.05986 | - | 0.27394 | 0.00893 |
| **CXCL13** | - | NS | NS | - | NS | NS |
| **CXCL9** | - | 0.09633 | 0.04795 | - | 0.18378 | NS |
| **FLRT2** | - | NS | NS | - | NS | NS |
| **GFAP** | - | 0.00350 | NS | - | 0.18602 | 0.12661 |
| **IL12B** | - | -0.07726 | -0.04695 | - | -0.43279 | -0.26344 |
| **MOG** | - | -0.08281 | -0.09053 | - | NS | NS |
| **NEFL** | - | 0.09751 | 0.00694 | - | 0.45991 | 0.08793 |
| **OPG** | - | NS | 0.00385 | - | NS | NS |
| **OPN** | - | NS | NS | - | NS | NS |
| **PRTG** | - | -0.11618 | -0.05996 | - | -0.72771 | -0.42898 |
| **SERPINA9** | - | -0.00412 | -0.00443 | - | -0.06799 | -0.05539 |
| **TNFRSF10A** | - | -0.00070 | NS | - | NS | NS |
| **TNFSF13B** | - | 0.10373 | 0.07722 | - | 0.48581 | 0.32122 |
| **VCAN** | - | -0.16151 | -0.10457 | - | -0.77226 | -0.56348 |

1. Please refer to Table 1 for description of clinical features.
2. Please refer to eTable 1 for description of protein features.

Abbreviations: NS = not selected by LASSO due to zero coefficient.

**eTable 5.** Feature coefficient of LASSO models predicting PROMIS.

| | PROMIS Binary | | | PROMIS Continuous | | |
|---|---|---|---|---|---|---|
| | Clinical Profile | Serum Protein Profile | Clinical Profile + Serum Protein Profile | Clinical Profile | Serum Protein Profile | Clinical Profile + Serum Protein Profile |
| | Clinical Features [1] | | | | | |
| **Age** | -0.00355 | - | NS | -0.28044 | - | -0.15617 |
| **Sex** | NS | - | NS | NS | - | NS |
| **Subtype** | -0.33248 | - | -0.20698 | -4.11563 | - | -1.04391 |
| **Disease Duration** | -0.00576 | - | -0.00631 | NS | - | -0.03201 |
| **Race/Ethnicity** | NS | - | NS | NS | - | NS |
| **DMT Efficacy** | -0.02264 | - | -0.02395 | -0.92613 | - | NS |
| **PROMIS Time** | NS | - | NS | 0.00082 | - | 0.00030 |
| | Protein Features [2] | | | | | |
| **APLP1** | - | 0.22378 | 0.12854 | - | 5.61844 | 5.03634 |
| **CCL20** | - | 0.02047 | NS | - | 0.02698 | NS |
| **CD6** | - | NS | NS | - | 1.38090 | 1.46740 |
| **CDCP1** | - | -0.29921 | -0.23801 | - | -6.63995 | -4.95301 |
| **CNTN2** | - | -0.11284 | -0.00068 | - | NS | NS |
| **CXCL13** | - | 0.01151 | NS | - | NS | NS |
| **CXCL9** | - | -0.04335 | NS | - | NS | NS |
| **FLRT2** | - | -0.00257 | NS | - | NS | NS |
| **GFAP** | - | -0.04518 | -0.01921 | - | -0.70370 | -0.28375 |
| **IL12B** | - | 0.17673 | 0.11815 | - | 1.58181 | 1.16752 |
| **MOG** | - | NS | NS | - | NS | NS |
| **NEFL** | - | -0.17193 | -0.09609 | - | -5.82478 | -2.99074 |
| **OPG** | - | NS | NS | - | -0.82371 | -0.40026 |
| **OPN** | - | -0.10643 | -0.04977 | - | -0.09135 | NS |
| **PRTG** | - | 0.27276 | 0.06075 | - | 2.95886 | 1.77987 |
| **SERPINA9** | - | -0.04953 | -0.00818 | - | NS | NS |
| **TNFRSF10A** | - | 0.11088 | NS | - | -1.05103 | NS |
| **TNFSF13B** | - | -0.02354 | NS | - | -2.41012 | -1.71371 |
| **VCAN** | - | -0.01382 | NS | - | 3.06650 | NS |

1. Please refer to Table 1 for description of clinical features.
2. Please refer to eTable 1 for description of protein features.

Abbreviations: NS = not selected by LASSO due to zero coefficient.

**eTable 6.** Predictive performances of alternative machine learning models in predicting binary PDDS or PROMIS as compared to the best LASSO models.

| PDDS | LASSO | Random Forest | | | XGBoost | | | Support Vector Machine | | |
|---|---|---|---|---|---|---|---|---|---|---|
| | Clinical Profile + Serum Protein Profile | Clinical Profile | Serum Protein Profile | Clinical Profile + Serum Protein Profile | Clinical Profile | Serum Protein Profile | Clinical Profile + Serum Protein Profile | Clinical Profile | Serum Protein Profile | Clinical Profile + Serum Protein Profile |
| AUC (95% CI) | 0.91 (0.85, 0.97) | 0.77 (0.65, 0.89) | 0.79 (0.68, 0.89) | 0.84 (0.73, 0.94) | 0.76 (0.64, 0.89) | 0.75 (0.64, 0.87) | 0.90 (0.83, 0.97) | 0.70 (0.56, 0.84) | 0.75 (0.62, 0.89) | 0.85 (0.76, 0.95) |
| Sensitivity | 0.89 | 0.50 | 0.45 | 0.48 | 0.58 | 0.46 | 0.67 | 0.60 | 0.80 | 0.88 |
| Specificity | 0.86 | 0.85 | 0.86 | 0.89 | 0.84 | 0.83 | 0.86 | 0.81 | 0.82 | 0.85 |
| PPV | 0.42 | 0.42 | 0.47 | 0.63 | 0.37 | 0.32 | 0.42 | 0.16 | 0.21 | 0.37 |
| NPV | 0.99 | 0.89 | 0.84 | 0.81 | 0.93 | 0.90 | 0.94 | 0.97 | 0.99 | 0.99 |
| F1-score | 0.57 | 0.46 | 0.46 | 0.55 | 0.45 | 0.37 | 0.52 | 0.25 | 0.33 | 0.52 |
| PROMIS | LASSO | Random Forest | | | XGBoost | | | Support Vector Machine | | |
| | Clinical Profile + Serum Protein Profile | Clinical Profile | Serum Protein Profile | Clinical Profile + Serum Protein Profile | Clinical Profile | Serum Protein Profile | Clinical Profile + Serum Protein Profile | Clinical Profile | Serum Protein Profile | Clinical Profile + Serum Protein Profile |
| AUC (95% CI) | 0.90 (0.78, 1.00) | 0.67 (0.38, 0.97) | 0.80 (0.61, 0.98) | 0.83 (0.68, 0.87) | 0.63 (0.31, 0.96) | 0.40 (0.14, 0.66) | 0.81 (0.65, 0.97) | 0.74 (0.49, 0.98) | 0.73 (0.52, 0.93) | 0.90 (0.80, 1.00) |
| Sensitivity | 0.93 | 0.90 | 0.87 | 0.90 | 0.86 | 0.84 | 0.90 | 0.91 | 0.86 | 0.86 |
| Specificity | 0.57 | 0.43 | 0.33 | 0.43 | 0.25 | 0.17 | 0.38 | 0.75 | 0.50 | 0.50 |
| PPV | 0.90 | 0.87 | 0.87 | 0.87 | 0.81 | 0.68 | 0.84 | 0.97 | 0.97 | 0.97 |
| NPV | 0.67 | 0.50 | 0.33 | 0.50 | 0.33 | 0.33 | 0.50 | 0.50 | 0.17 | 0.17 |
| F1-score | 0.92 | 0.89 | 0.87 | 0.89 | 0.83 | 0.75 | 0.87 | 0.94 | 0.91 | 0.91 |

Abbreviations: AUC = area under the ROC curve, 95% CI = 95% confidence interval, PPV = positive predictive value, NPV = negative predictive value

**eTable 7.** Predictive performances across different stacking ensemble models in predicting binary PDDS or PROMIS.

| LASSO [1] | - | - | | Random Forest [2] | - | - |
|---|---|---|---|---|---|---|
| | **PDDS** | **PROMIS** | | | **PDDS** | **PROMIS** |
| **AUC (95% CI)** | 0.89 (0.82, 0.97) | 0.80 (0.63, 0.97) | | **AUC (95% CI)** | 0.84 (0.74, 0.95) | 0.86 (0.74, 0.98) |
| **Sensitivity** | 0.67 | 0.93 | | **Sensitivity** | 0.70 | 0.90 |
| **Specificity** | 0.86 | 0.44 | | **Specificity** | 0.85 | 0.38 |
| **PPV** | 0.42 | 0.84 | | **PPV** | 0.37 | 0.84 |
| **NPV** | 0.94 | 0.67 | | **NPV** | 0.96 | 0.50 |
| **F1-Score** | 0.52 | 0.88 | | **F1-Score** | 0.48 | 0.87 |

| XGBoost [3] | - | - | | SVM [4] | - | - |
|---|---|---|---|---|---|---|
| | **PDDS** | **PROMIS** | | | **PDDS** | **PROMIS** |
| **AUC (95% CI)** | 0.82 (0.72, 0.91) | 0.67 (0.44, 0.90) | | **AUC (95% CI)** | 0.68 (0.56, 0.79) | 0.69 (0.46, 0.91) |
| **Sensitivity** | 0.75 | 0.90 | | **Sensitivity** | 0.90 | 0.90 |
| **Specificity** | 0.84 | 0.38 | | **Specificity** | 0.43 | 0.43 |
| **PPV** | 0.32 | 0.84 | | **PPV** | 0.87 | 0.87 |
| **NPV** | 0.97 | 0.50 | | **NPV** | 0.50 | 0.50 |
| **F1-Score** | 0.44 | 0.87 | | **F1-Score** | 0.89 | 0.89 |

1. Stacking ensemble learning using LASSO as the second level model (Refer to Methods in the main text and Figure 1C)
2. Stacking ensemble learning using Random Forest as the second level model
3. Stacking ensemble learning using XGBoost as the second level model
4. Stacking ensemble learning using SVM as the second level model

Abbreviations: AUC = area under the ROC curve, 95% CI = 95% confidence interval, PPV = positive predictive value, NPV = negative predictive value
